# The SARS-CoV-2 pandemic course in Saudi Arabia: A dynamic epidemiological model

**DOI:** 10.1101/2020.06.01.20119800

**Authors:** Abdullah Murhaf Al-Khani, Mohamed Abdelghafour Khalifa, Abdulrahman AlMazrou, Nazmus Saquib

## Abstract

**Objective:** Saudi Arabia ranks second in the number of coronavirus disease 2019 (COVID-19) cases in the Eastern Mediterranean region. It houses the two most sacred religious places for Muslims: Mecca and Medina. It is important to know what the trend in case numbers will be in the next 4–6 months, especially during the Hajj pilgrimage season.

**Methods:** Epidemiological data on COVID-19 were obtained from the Saudi Arabian Ministry of Health, the World Health Organization, and the Humanitarian Data Exchange. A susceptible-exposed-infectious-recovered (SEIR) prediction model was constructed to predict the trend in COVID-19 in Saudi Arabia in the next 6 months.

**Findings:** The model predicts that the number of active cases will peak by 20 May 2020. The cumulative infected cases are predicted to reach 59,663 at that time. The total number of infected individuals is estimated reach to 102,647 by the end of the pandemic.

**Conclusion:** Our estimates show that by the time the Hajj season commences in Saudi Arabia, the pandemic will be in the midst of its deceleration phase (phase 3). This information will likely be useful to policymakers in their management of the outbreak.

## 1. Introduction

Since late December 2019, the world has been experiencing a rapidly spreading, highly infectious virus, the severe acute respiratory syndrome coronavirus 2 (SARS-CoV-2). The World Health Organization (WHO) declared this new disease a “Public Health Emergency of International Concern” on 30 January 2020 and then as a “Global Pandemic” 40 days later^(1)^. Current knowledge suggests that respiratory droplets are the primary mode of transmission. Less than 50% of all active coronavirus disease 2019 (COVID-19) cases are asymptomatic, which is lower than that of influenza (56–80% asymptomatic)^(2)^.

Within the Eastern Mediterranean region, the Kingdom of Saudi Arabia ranks second only to the Islamic Republic of Iran in total number of infected and active cases^(3)^. Saudi Arabia has a population of 34 million, the majority (37%) of whom dwell in several major cities: Riyadh, Mecca, Jeddah, Dammam, and Medina^(4)^. The majority of COVID-19 cases are concentrated within these cities. As of 28 May 2020, there were 80,185 total cases in Saudi Arabia, 54,553 recoveries and 441 deaths^(5)^. The healthcare system of Saudi Arabia is no stranger to coronavirus outbreaks, having dealt before with local outbreaks of Middle East respiratory syndrome coronavirus (MERS-CoV)^(6)^.

The first COVID-19 case (n = 1) in Saudi Arabia was detected in the Eastern region (i.e., Qatif) on March 2nd; it was an individual who had traveled to the endemic region of Iran.

The Qatif area was put on lockdown within three days of the first case. The government took decisive measures to control potential outbreaks by cancelling festivals and events, suspending e-Visa entry, and halting all domestic travel by March 7^th^. The following day, all educational institutions were closed. Public gatherings and weddings were banned a week later. On March 15^th^, all arriving and departing international flights were cancelled, followed on March 17^th^ by the prohibition of daily prayers in mosques, including the weekly Friday prayer. Transportation between cities was banned soon after on March 21^st(7)^. To mitigate virus transmissions that occur from gatherings, Saudi authorities have enforced a 24-hour curfew in major cities (Riyadh, Medina, Jeddah, Mecca, and Dammam) since April 6^th^, while the rest of the country has been under limited curfew (from 3 pm to 6 am) since April 7^th(8)^. It announced a 5-day nationwide 24-hour curfew from May 23^rd^ to May 27^th^ to curb the tide of the epidemic^(9)^.

Saudi Arabia houses the two most sacred religious places for Muslims: Mecca and Medina. They make pilgrimage to these cities either at a fixed time (i.e., Hajj) or at any time (i.e., Umrah) in a calendar year. During the current pandemic, Saudi authorities have stopped Umrah pilgrimage, effectively reducing imported SARS-CoV-2 cases^(10)^. Furthermore, Hajj has been put on hold this year due to the overwhelming infection rate of SARS-CoV-2^(11)^. Since 2009, Saudi Arabia has faced multiple outbreaks during Hajj seasons, including the H1N1 pandemic^(12)^, multiple local MERS-CoV outbreaks^(13)^, and Ebola and Zika viruses from African pilgrims^(14, 15)^. The literature has suggested Hajj pilgrims played a key role in the dispersion of the 1957 flu pandemic^(16)^.

Infectious disease prediction models are tools that use available data about the status and progress of an infectious outbreak to predict its future course^(17)^. In its most basic form, an infectious disease prediction model is constructed of three essential compartments: 1) susceptible (S), 2) infected (I), and 3) recovered (R), commonly abbreviated as the SIR model. The addition of a fourth compartment, i.e., exposed (E), increases the prediction capability of the resultant model (i.e., SEIR), which is now being widely used^(18)^.

There are practical implications for predicting the epidemiological trajectory of the COVID-19 in Saudi Arabia as the country is still in the midst of this outbreak. A prediction can inform the country’s decision makers and help them make prudent decisions about the continued management of the outbreak. The Saudi healthcare system can also utilize this information to adapt to changing health needs. Meanwhile, we can hope to maintain a controlled situation until vaccines and treatments are deployed.

We name our prediction model KSA-CoV-19, and with it, aim to find the following: 1) the anticipated epidemic curve of SARS-CoV-2 in Saudi Arabia, 2) the peak, the end, and the number of COVID-19 cases associated with the curve, and 3) the timing of upcoming Hajj 2020 (July 28^th^ –August 2^nd^) in relation to the anticipated epidemic curve.

## 2. Material and methods

### 2.1. Epidemiological data

The data used to generate our KSA-CoV-19 Model were obtained from three different sources: 1) the Saudi Arabian Ministry of Health^(5)^, 2) the WHO^(3)^, and 3) the Humanitarian Data Exchange^(19)^. The three databases were cross-checked against each other for accuracy and as a sensitivity measure. While all data sources provided an identical reporting of the number of confirmed, recovered and death cases, none of them provided any daily updates in respect to the number of quarantined (i.e., exposed) cases. A recent report from the Centers for Disease Control and Prevention found that for 10 positive cases, 445 close-contact individuals were identified^(20)^. We used this information to estimate the number of the exposed compartment in the model.

### 2.2. Epidemiological model compartments

A deterministic compartmental model (DCM) with a susceptible-exposed-infectious-recovered (SEIR) disease class was chosen to be implemented in the KSA-CoV-19 Model. For this model to work, a number of assumptions/variables needed to be inputted. Firstly, there were parameters that characterize the disease: 1) the basic reproduction number (R0), 2) the duration of the exposed state (e.dur), 3) the duration of the infectious state (i.dur), and 4) the case fatality rate (cfr). Secondly, the population parameters at the start of the modeling were also inputted: 1) number of susceptible (s.num), 2) the number of exposed (e.num), 3) the number of infected (i.num), and 4) the number of recovered (r.num). Building on the work of Peng et al., a new compartment (P) was included in our model that conveyed the increased level of public health awareness, such as widespread face masks, strict social distancing, and the locking-down of cities^(21)^. We assumed that the susceptible population(S) was declining at a steady rate as a result of a positive protection rate (α); the number of those protected (P compartment, p.num) is removed from the susceptible pool (s.num).

In this paper, we present four models: KSA-CoV-19 Model, two variants of KSA-CoV-19 Model, and a Natural Course model (Table 1). The parameters chosen for these four models are justified below.

**Table 1.** Different parameters used to generate the epidemiological models of the COVID-19.

| Parameters | Natural Course Model | KSA-CoV-19 Model | Model 2 | Model 3 |
| --- | --- | --- | --- | --- |
| R0 | 5.7 | 5.7 | 5.7 | 5.7 |
| e.dur | 6.4 days | 6.4 days | 6.4 days | 6.4 days |
| i.dur | 7 days | 7 days | 7 days | 3 days |
| cfr | 0.86% | 0.86% | 0.86% | 0.86% |
| $\alpha$ | 0 | 0.023 | 0.18 | 0.023 |
| s.num | 33,999,959 |  |  |  |
| e.num | 40 |  |  |  |
| i.num | 1 |  |  |  |
| r.num | 0 |  |  |  |
| p.num | 0 |  |  |  |

R0: basic reproduction number; e.dur: duration of the exposed state; i.dur: duration of the infectious state; cfr: case fatality rate; $\alpha$ : protection rate; s.num: number of susceptible; e.num: number of exposed; i.num: number of infected; r.num: number of recovered; p.num: number of protected

### 2.3. Parameter estimation

We assumed that our model is based on a closed system (i.e., S+P+E+I+R = 34 million, the total population of Saudi Arabia)^(4, 22)^, and that deaths in the population are solely due to COVID-19. In our model, we did not use parameters related to the birth and death rate among the susceptible, exposed, infectious, and recovered, in line with the methodology used in similar studies^(22)^.

We chose March 2^nd^, the day on which the first positive case (i.e., diagnosed) of COVID-19 was documented, to be the starting date of our prediction model. In the absence of definitive data, we assumed that the first diagnosed case exposed the average 40 people.

In their report, Sanche et al. estimated the median R0 value of SARS-CoV-2 to be 5.7 (95% CI 3.8–8.9)^(23)^. Thus, the KSA-CoV-19 Model integrated an R0 value of 5.7. Furthermore, other reports found that the average incubation period of SARS-CoV-2 was 6.4 days^(24)^, which we have used as the e.dur in our model.

Woelfel et al. estimated an average duration of infection (i.dur) of 3 days or 7 days^(25)^. Therefore, a conservative period of 7 days was inputted as the duration of the infected state(i.dur) in the KSA-CoV-19 Model (Table 1). We used a 3-day duration to generate another model (Model 3, Table 1). The aforementioned parameter estimates (i.e., e.dur, i.dur) have been previously used in other SEIR models^(22)^.

In regards to the death rate among those infected, although the WHO states that the global mortality rate of COVID-19 is approximately 3.4%^(26)^, in Saudi Arabia, a rate of 0.86% was calculated by averaging the death rate from when the first death case was recorded until 9 April 2020. We used this rate (0.86%) in all four models that we present.

After trial and error, we chose an α (protection rate) value of 0.023 for our model. This value produced a prediction line that most closely fit the observed cases in Saudi Arabia so far. As a sensitivity measure, we used values reported by other studies that have utilized a similar methodology. For example, in their original report, Peng et al.^(21)^ used a value of 0.18, so we used that to generate another model (Model 2, Table 1).

Finally, we generated a model that simulated the natural course or “free fall” of the SARS-CoV-2 virus in Saudi Arabia. This model assumes that people were mixing at random and that no preventive measures were taken to halt the progression of the infection (Natural Course Model, Table 1).

### 2.4. Statistical software

The analytic component of this paper was carried out using the EpiModel^(27)^ package in the R statistical programming language, version 3.6.3 (R Foundation for Statistical Computing). Microsoft Excel 2016 was used for data storage and management.

## 3. Results

In this paper, we generated four different models that provided an estimation for the COVID-19 course in Saudi Arabia. A summary of the models is shown in Fig. 1.

**Fig. 1.**
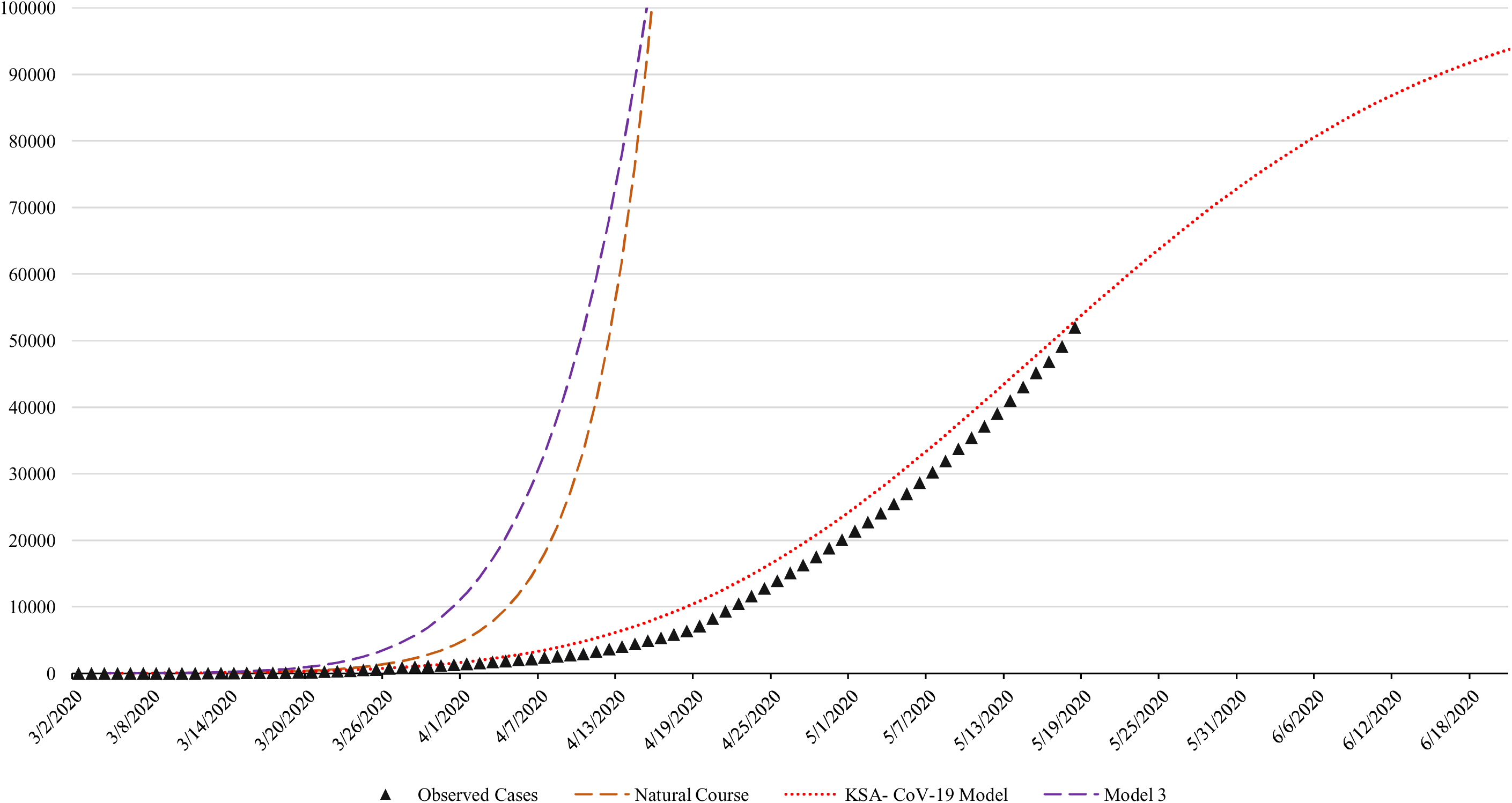
The trend of COVID-19 in Saudi Arabia. Three different models are plotted to predict the growth of cases.

The Natural Course Model shows a hypothetical, worst-case scenario in which no preventive measures are taken. The ascending phase (phase 2) would start on April 21^st^, and the peak would be reached by May 18^th^. At the peak, the cumulative case count would be 23,351,139 (68.6% of the total population). The descending phase (phase 3) would conclude approximately by July 1^st^. A total of 33,570,820 would be infected by that time.

Out of all the models generated, the KSA-CoV-19 Model most closely fit the observed data. This model takes into account the preventive measures that are to be implemented to halt the progression of the disease, introduced into the model as a factor of α. In this model, the second (acceleration) phase lasts a period of 54 days from March 27^th^ till the peak on May 20^th^. The number of cumulative cases at the peak is 59,663 cases. The third (deceleration) phase will finish by August 14^th^, with a total of 102,589 infected individuals by that time. The 2020 Hajj season will coincide with the deceleration leg of this pandemic curve. The total number of infected individuals is estimated to reach 102,647 by the end of the pandemic. Fig. 2 shows the predicted number of active COVID-19 cases for the KSA-CoV-19 Model.

**Fig. 2.**
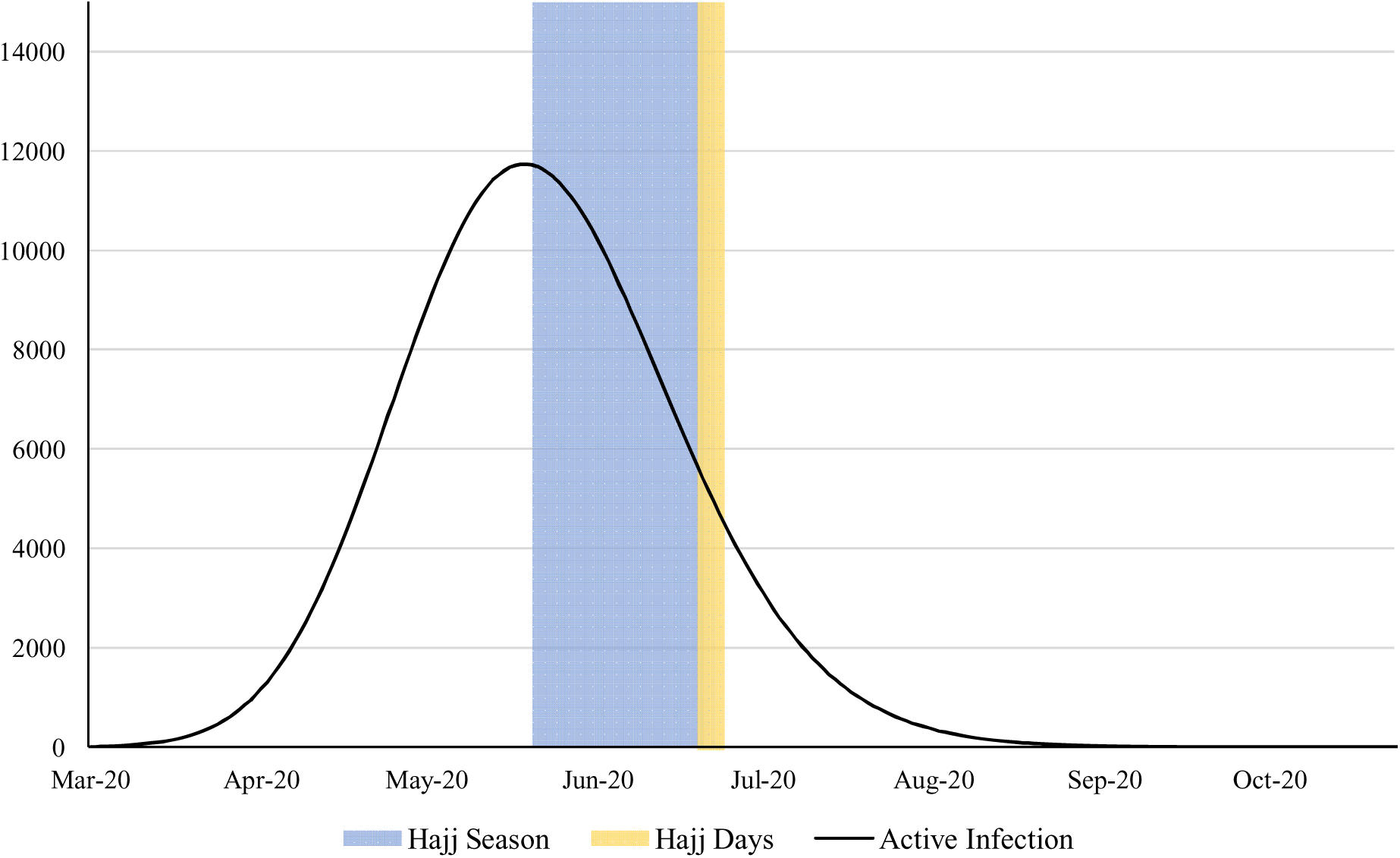
The predicted number of actively infected cases in Saudi Arabia according to the KSA-CoV-19 Model. The duration of the Hajj Season is shown in blue.

Using an α (protection rate) value of 0.18, similar to that used in the model constructed by Peng et al., resulted in a quick resolution of the pandemic in Model 2, which predicted a peak on March 15^th^ and a total of 78 infected individuals by that time. The deceleration (third) phase finishes by April 19^th^, with the number of infected people not exceeding 122 individuals. The curve of this model was omitted from Fig. 1 due to its minuteness.

Model 3 implemented the same parameters used in the KSA-CoV-19 Model, changing only the duration of the infected state (i.dur) from 7 days to 3 days. This model closely resembles the Natural Course model, albeit with lower magnitude. A peak is reached by May 13^th^, with 1,086,332 total infections. The end of the third phase is reached by June 30^th^, and the number of infected is projected to reach 1,997,404.

## 4. Discussion

The rapid evolution of the COVID-19 situation worldwide and in Saudi Arabia makes it extremely challenging for healthcare systems to adapt. This paper predicts the course of the pandemic in the coming months in Saudi Arabia. We predict that the effects of the COVID-19 will have peaked during the third week of May 2020.

Availability of epidemiological modeling of the novel corona virus in the Middle East is scarce. Indian researchers recently predicted that COVID-19 cases would peak between the third and fourth weeks of April 2020 in India, when they would reach 68,978 confirmed cases^(28)^. Recent reports have stated that the current forecasts project a continuing increase with large uncertainty^(29)^.

Our mathematical model has several strengths. For example, data used in the modeling were obtained from three different sources and checked for accuracy. Additionally, this model incorporated a susceptible-exposed-infectious-recovered (SEIR) mathematical model, which is more favorable than a susceptible-infectious-recovered (SIR) one.

On the other hand, one limitation of this study is that the accuracy of the predictions only lasts for a few weeks to months due to the erratic behavior of the current corona virus. Similarly, the accuracy of this prediction is highly correlated with how accurately and effectively new cases are recorded; it is vital to note that the number of identified infected cases will largely depend on the implemented testing strategy (i.e., how many tests are done in the population). As of May 28^th^, Saudi Arabia had done 770,696 tests, which is equal to approximately one test per 45 people^(5)^. Additionally, our KSA-CoV-19 Model overestimates the number of recovered cases, but the number of recovered cases in Saudi Arabia has been increasing rapidly. For instance, in a period between 5 May and 15 May 2020, the number of recoveries increased 4-fold, and by May 28^th^, the number of recovered cases had surpassed the number of active cases by a large margin^(5)^. Finally, despite utilizing previously published estimates for the average incubation period (e.dur) and average infection duration (i.dur), the model still undershoots the amount of active infection due to the rapid flow from the infected (I) compartment to the recovered (R) compartment of the SEIR model.

## 5. Conclusion

In conclusion, this KSA-CoV-19 Model is one of the few and early prediction models in Saudi Arabia and the Middle East. Our estimates show that by the time the Hajj season commences in Saudi Arabia, the pandemic will be in the midst of its deceleration phase (phase 3), during which the danger of the pandemic will be declining, but exceptional care should be taken to avoid a resurgence. Strict adherence to the current control measures is essential to maintain the predicted pattern, and caution should be taken when easing these measures because deviation may adversely alter the predicted course. The final decision on whether to hold the Hajj pilgrimage this year should take the findings of this study into consideration.

## Data Availability

The data that support the findings of this study are available from the corresponding author upon reasonable request.

## Abbreviations

cfr: mortality proportion among the infected
CDC: Centers for Disease Control and Prevention
COVID-19: coronavirus disease 2019
DCM: deterministic compartmental model
e.dur: duration of the exposed state
e.num: number of exposed
i.dur: duration of the infectious state
i.num: number of infected
MERS-CoV: Middle East respiratory syndrome coronavirus
p.num: number of protected
r.num: number of recovered
R0: basic reproduction number s.num: number of susceptible
SARS-CoV 2: severe acute respiratory syndrome coronavirus 2
SEIR: susceptible, exposed, infectious and recovered infectious disease prediction model
SIR: susceptible, infectious and recovered infectious disease prediction model

## Acknowledgement

The authors thank Ms. Erin Strotheide for her editorial contributions to this manuscript.

## Author contributions

AMA conceived of the study, designed and programmed the model, interpreted the results, and prepared the original draft. MAK prepared the original draft. AA consulted on the analyses and reviewed and edited the final manuscript. NS consulted on the analyses and reviewed and edited the final manuscript.

